# Fingerprint: An AI-based method for detection of mislabeled CT studies in clinical trials

**DOI:** 10.1101/2022.12.21.22283802

**Authors:** Lars Edenbrandt, Olof Enqvist, Tony Gillberg

## Abstract

The manual step of replacing the identification number of each subject in clinical trials with the correct trial identification number is critical. Mislabeling studies may lead to excluding subjects or visits or to false study results. This article presents an automated AI-based method to detect if a pair of de-identified CT studies has been obtained from the same subject.

**Approach:** An automated segmentation of bones in CT is performed using Organ Finder (SliceVault AB, Malmö, Sweden). Corresponding bones from the two images are aligned using Iterative closest point. The percentage of vertices in the smaller mesh with residuals below 1.5 mm is used as a measure of similarity, the *anatomical match*. If the anatomical match is less than 80% for the left and right hip bone and the left and right scapula, the two CT studies are classified as being obtained from different subjects. The Fingerprint method was tested on a group of 58 patients, who each had two CT studies obtained at different occasions. From the 116 CT studies 6,612 unique pairs of CT from different patients and 58 pairs from the same patient could be selected.

**Results:** The Fingerprint method classified all 6,612 pairs of CT studies from different patients correctly (sensitivity 100%; 95% confidence interval 99-100%) and all 58 pairs of CT studies from the same patients correctly (specificity 100%; 95% confidence interval 94-100%).

**Conclusion:** This study shows how an AI-based method can be used to accurately detect if a pair of de-identified CT studies has been obtained from two different subjects.

## Introduction

In clinical trials, diagnostic images are commonly de-identified and uploaded to a central repository for further analysis. The manual step of replacing the identification number of each subject with the correct trial identification number is critical. Mislabeling studies may lead to excluding subjects or visits or to false study results. In some situations, detecting that two CTs are obtained from different subjects is easy by visual inspection, for example if the subjects differ in size, gender, or if one subject has a hip prosthesis, but it can easily be overlooked especially as mistakes are rare and the focus of the image reader is on other tasks such as tumor detection. Artificial intelligence (AI)-based methods are well suited to handle this issue, tirelessly looking for the unlikely mistake. This article presents an automated AI-based method to detect if a pair of de-identified CT studies has been obtained from the same subject or not.

## Methods

### Patients

#### Training set

The Fingerprint method was developed using a group of lymphoma patients who had undergone more than one PET/CT examination for the purpose of treatment response evaluation. Only the CT images were used in this study. The training set consisted of 8 female and 19 male patients with a median age of 34 years (range 15 to 73 years).

PET/CT data were obtained using an integrated PET/CT scanner (Siemens Biograph 64 Truepoint). A low-dose CT scan (64-slice helical, 120 kV, 30 mAs, 512 × 512 matrix) was obtained covering the base of the skull to the mid-thigh. Slice thickness and spacing between slices were 3 mm.

There was a total of 63 studies from which 1,953 unique study pairs could be selected. Out of these, 46 pairs consisted of CT studies from the same patient obtained at different occasions and the remaining 1,907 pairs consisted of CT studies from different patients. Ethical approval was granted by the ethics committee at Gothenburg University (#295-08).

#### Test set

The Fingerprint method was evaluated using a publicly available dataset of 58 female patients with a median age of 52 years (range 22 to 83 years) [1-2].

PET/CT data were obtained using integrated PET/CT scanners (Discovery LS, ST, and STE, GE Medical Systems, Gemini TF, Philips Medical Systems, Biograph 6, 40, 64 and Sensation 16 Siemens). A CT scan (120-140 kV, 42-398 mAs, 512 × 512 matrix) was obtained covering the base of the skull to the mid-thigh. Slice thickness and spacing between slices were 2-4.25 mm.

There were two CT studies for each patient. From the 116 studies, 6,670 unique study pairs were selected. Out of these pairs, 58 consisted of CT studies from the same patient obtained at different occasions and the remaining 6,612 pairs consisted of CT studies from different patients.

### The Fingerprint Method

The first step of the Fingerprint method is an automated organ segmentation. The AI-tool Organ Finder (SliceVault AB, Malmö, Sweden) is used to segment organs in the two CT studies to be compared [3]. Organ Finder is based on a convolutional neural network (CNN) trained on 1,151 CT studies with manual organ segmentations. This CNN segments 22 different organs and four of those, the left and right hip bone and the left and right scapula, are used in the Fingerprint method (Figure 1).

**Figure 1.**
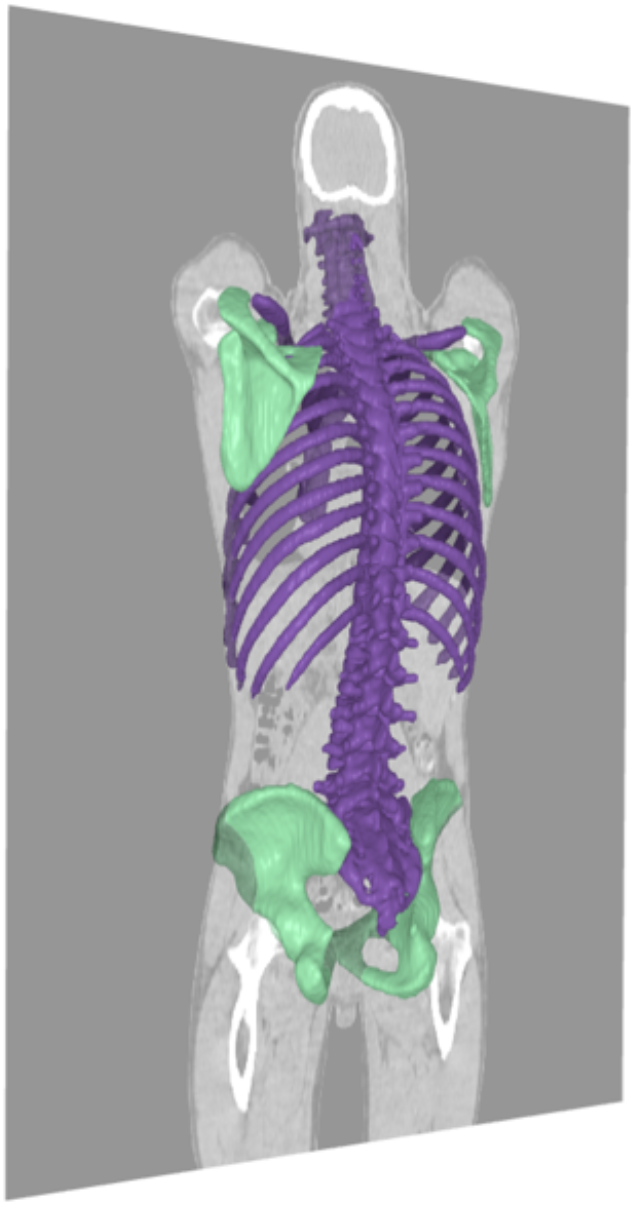
Bone segmentation from Organ Finder. The four green bones are used in the Fingerprint method.

The second step is to calculate an anatomical match measure describing the similarity between bones segmented from the two CT studies to be compared. SurfaceNets by Gibson is used to extract meshes describing the bone surfaces [4]. The meshes from the two studies are aligned using Iterative closest point (ICP) [5] to put them in a common coordinate system (Figure 2). For each vertex in the smaller mesh the closest distance to the other mesh is computed and the percentage of the vertices with residuals below a predefined threshold is used as a measure of similarity, the *anatomical match*.

**Figure 2.**
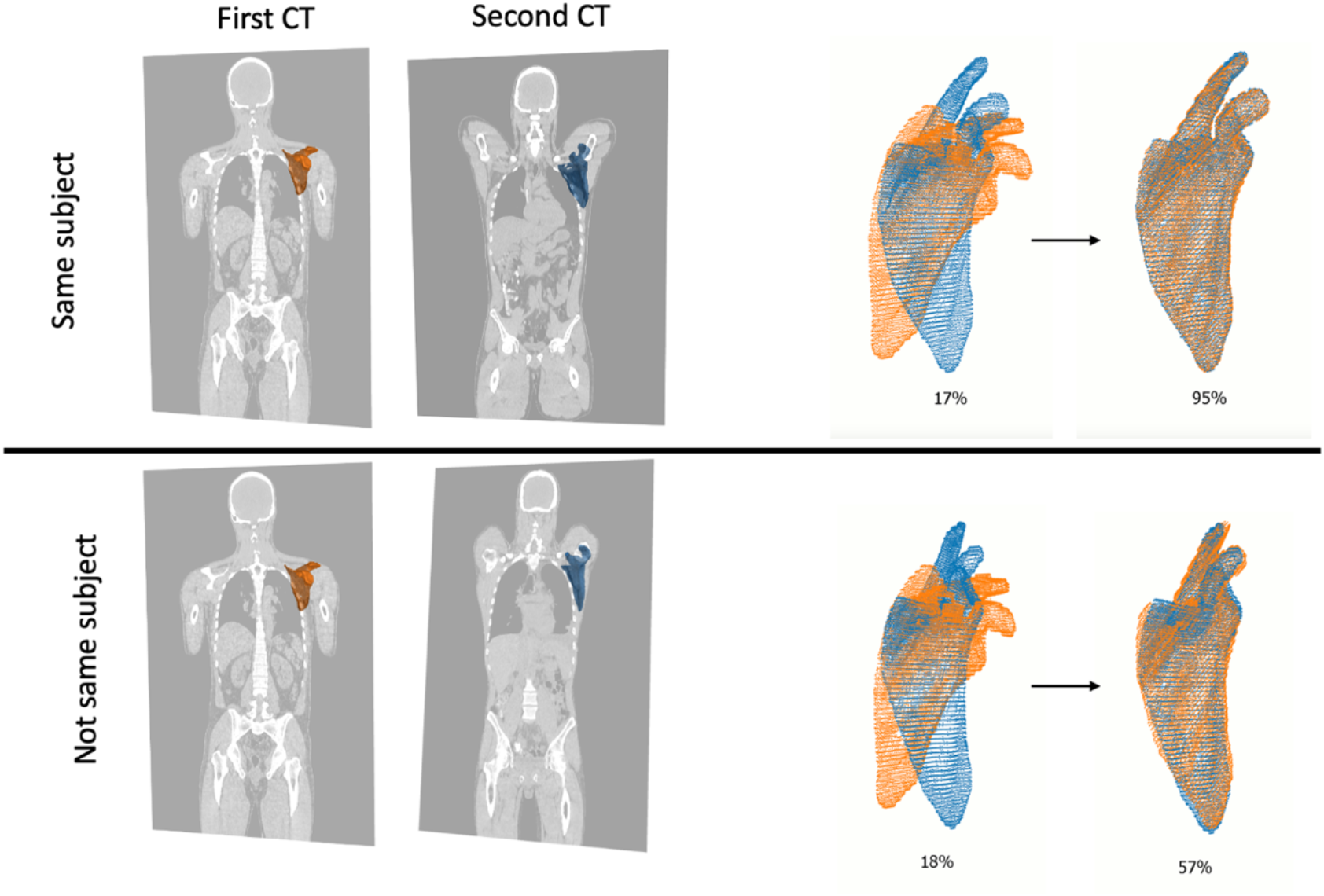
Alignment of left scapula from two CT studies. The two studies above are from the same subject and the two studies below are from different subjects. The bones are aligned in 3D and the final anatomical match is 95% for the pair from the same subject and 57% for the pair from different subjects.

Different thresholds were evaluated on the training set before selecting 1.5 mm. The anatomical match for each image pair in the training set, is shown in Figure 3. The separation between pairs of CT studies from the same and different patients was good both for hip bones and for scapulae.

**Figure 3.**
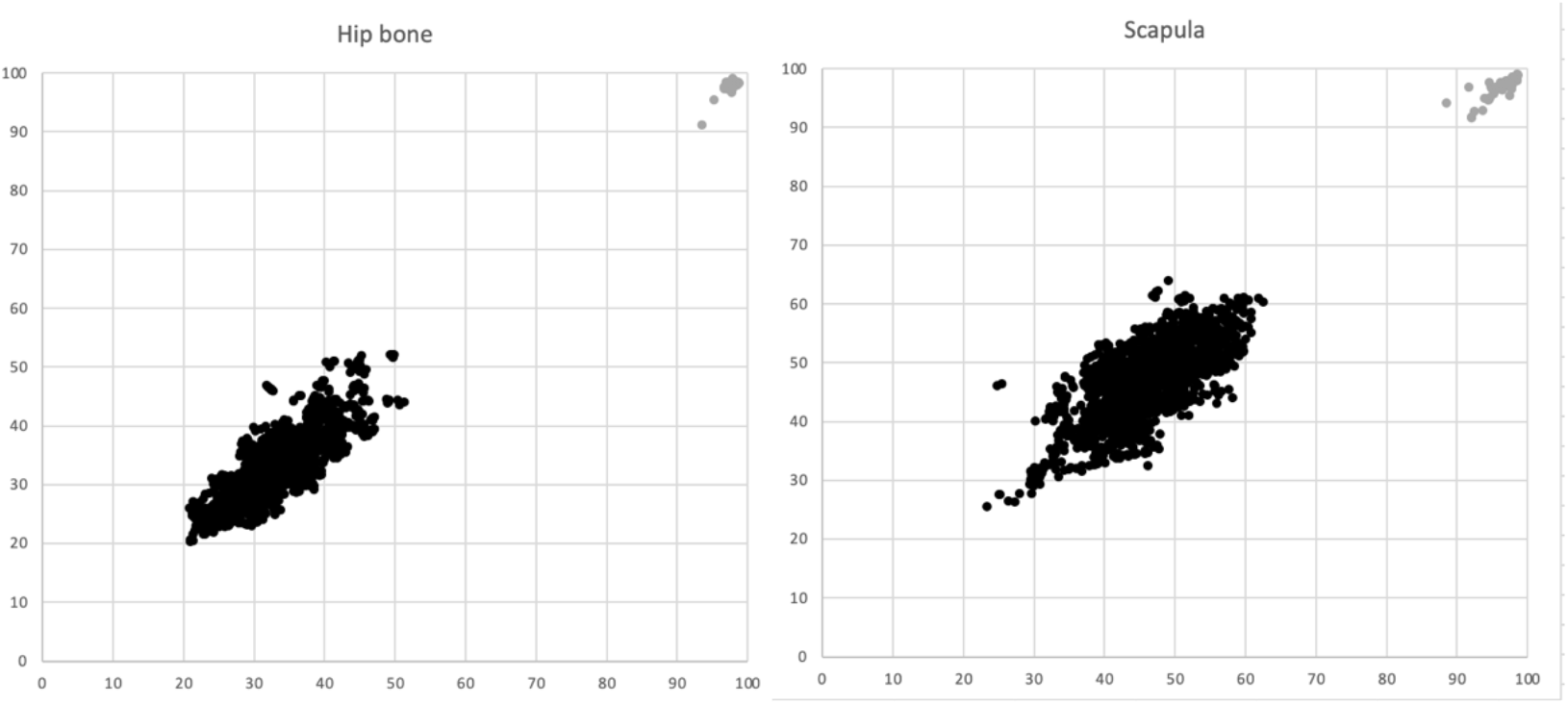
Anatomical match for the left (Y axis) and right (X axis) sided hip bone and scapula. Grey dots represent pairs of CT studies from the same subject (n=46) and black dots represent pairs of CT studies from different subjects (n=1,907).

The following criterion, based on the training set results, was implemented in the Fingerprint method. If the anatomical match is less than 80% for all four bones, the two CT studies are classified as being obtained from different subjects. This criterion correctly classified all pairs in the training set.

### Statistical methods

The Clopper-Pearson (exact) method was used to calculate the 95% confidence intervals (CI) for a binomial probability.

## Results

The Fingerprint method classified all 6,612 (sensitivity 100% CI 99-100%) pairs of CT studies from different subjects correctly and all 58 pairs of CT studies from the same subjects correctly (specificity 100% CI 94-100%). Figure 4 shows the data points separately for hip bone and scapula.

**Figure 4.**
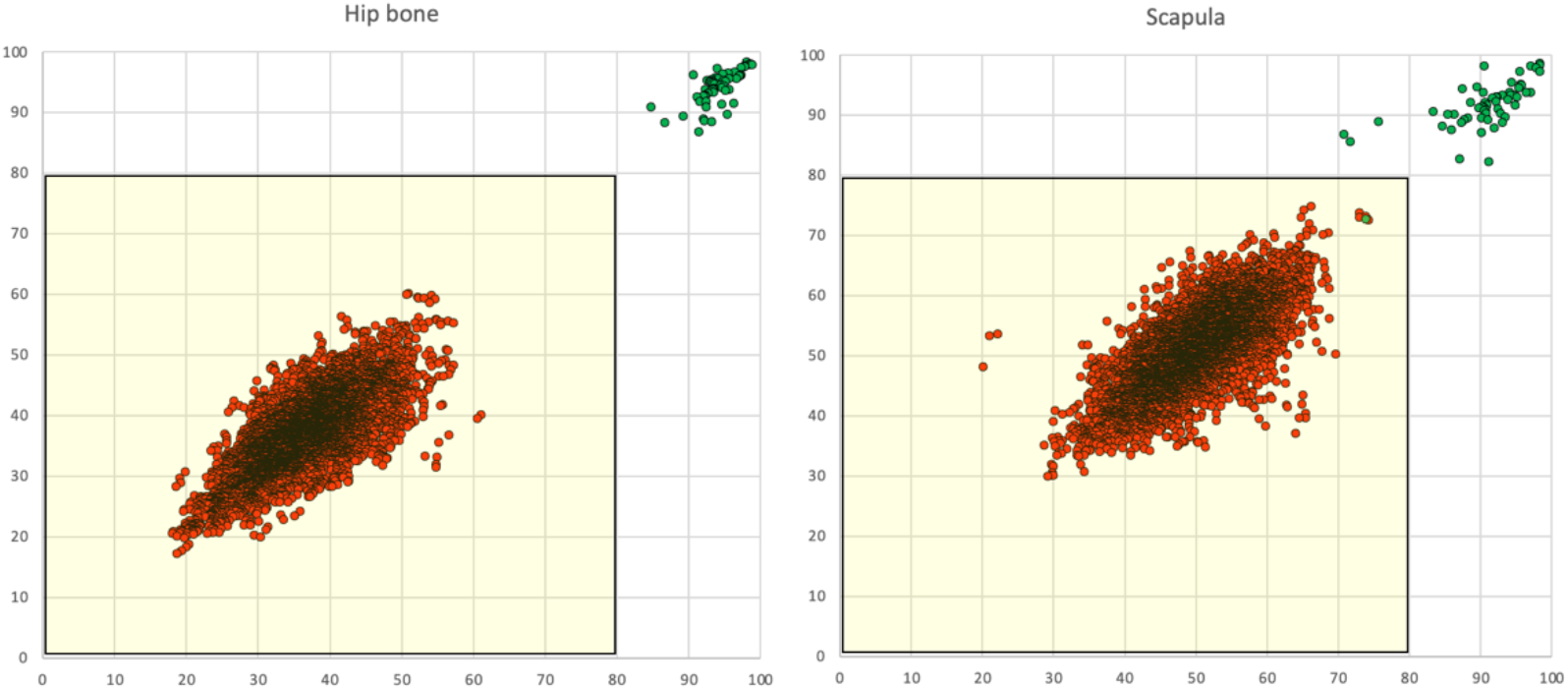
Anatomical match for the left (Y axis) and right (X axis) sided hip bone and scapula, respectively. Green dots represent pairs of CT studies from the same subject (n=58) and red dots represent pairs of CT studies from different subjects (n=6,612).

## Discussion

This study shows that mislabeled CT studies in clinical trials can be detected by comparing the shape of bone structures. The shape and size of a hip bone and scapula mimic the feature of a human fingerprint in being unique for every individual, but our CT-based Fingerprint method is not intended to identify individuals with high degree of certainty, but to alert clinical trial staff that a CT study may be mislabeled.

The Organ Finder tool was used to segment the hip bones and scapulae of the 63 CT studies. The segmentations showed to be very accurate. This is in agreement with the results from the paper describing Organ Finder in which the Sørensen-Dice index was 0.97 and 0.94 respectively for these to bones [3]. A good organ segmentation is essential for the Fingerprint method to be accurate.

The CT studies of the test set were obtained with scanners from different vendors. The protocols used also differed with for example slice thickness ranging from 2 to 4.25 mm. The good results despite this variation indicate that the Fingerprint method can be useful in clinical trials where this type of variation is common.

A limitation of the present study is the relatively small number of same-subject CT pairs. In the real application, this type of pairs will be much more common than pairs from different subjects, so it is crucial to keep the number of false alarms down. Our results indicate that our current thresholds would result in few false alarms, but that has to be evaluated on a larger material.

The training set included predominantly male patients and the test set only female patients. A more well-balanced training set would be preferrable before testing the method in a larger population.

Future work will also include adding more bones for analysis. At present the Organ Finder cannot segment separate vertebrae, but when that is included the Fingerprint method can be updated.

Quality checks of images will become more and more important in clinical trials. Good image quality is vital for reliable results and routines to check images are essential. We have recently shown that it is feasible to use AI-based methods to automatically perform a quality assessment with a very high accuracy [6]. The AI-based method in that study checked CT studies, both regarding the parts of the body included (i.e., head, chest, abdomen, pelvis), and other image features (i.e., presence of hip prosthesis, intravenous contrast and oral contrast). The Fingerprint method developed in this study is another example of an AI method that can be of value in future clinical trials.

In conclusion, this study shows how the AI-based Fingerprint method can be used to accurately detect if pairs of de-identified CT studies have been obtained from different subjects.

## Data Availability

The test group is a publicly available dataset. The training group is not publicly available for ethical reasons.

https://wiki.cancerimagingarchive.net/pages/viewpage.action?pageId=30671268

